# Comparison of epidemiological characteristics of COVID-19 patients in Vietnam

**DOI:** 10.1101/2020.06.03.20121467

**Authors:** Toan Ha, Gualberto Ruaño, Lewis Judy

**Author notes:** **Corresponding Author** Toan Ha, School of Medicine, University of Connecticut, 263 Farmington Avenue, Farmington, CT 06030.

## Abstract

**Background:** There are limited data on COVID-19 patients in Vietnam. The paper examined and compared epidemiological characteristics of COVID-19 patients in Vietnam

**Method:** The data was obtained using publicly available information from the official website of Vietnam Ministry of Health covering a period of 01/23/2020 to 05/27/2020. T-test, Chi-square test and Fisher’s Exact test were utilized to compare characteristics of COVID-19 patients between under-treatment and discharge groups and between overseas and non-overseas travel groups.

**Results:** Vietnam had a total of 327 cases of COVID-19 as of May 27, 2020. The median age of patients was 30 years (ranging from 3 months to 88 years). About 68% of patients (n=223) had acquired the disease from overseas while 32% were infected from local transmission. Among those infected from local transmission, 66% were women. Men were more likely than women to be infected with COVID-19 from overseas (p<0.001). Younger patients were significantly associated with international travel (p=.001). While patients in the South reported highest levels of overseas travel history (77.9%), those (100%) in the Central reported the highest level of being discharged (p<0.001). Women (54.7%) had a higher rate of discharge compared to men (45.3%) [p <0.001]. Nearly 86% have recovered and discharged from hospitals. There has been no reported fatality.

**Conclusions:** A majority of COVID-19 cases in Vietnam were acquired overseas. A significantly higher number of women than men were infected inside the country calling for further research about gender disparities in the fight against COVID-19 in Vietnam.

## Introduction

The outbreak of COVID-19 began in China on December 2019 and has quickly spread all over the world [1]. The World Health Organization (WHO) declared Covid-19 a pandemic on March 11, 2020 [2]. As of May 27, the total number of confirmed cases was 5,549,131 and 348,224 deaths were reported worldwide [3].

Vietnam, a country with 97 million people, had its first Covid-19 case in January 23, 2020 [4]. As of May 27, Vietnam has had a total of 327 COVID-19 confirmed cases with no fatalities [5]. The COVID-19 pandemic in Vietnam can be divided into two waves [6]. The first wave included 23 patients, which mainly related to the imported cases from China. These patients were successfully treated and discharged from hospitals [5]. The second wave started with cases imported from Europe mainly from the United Kingdom and spread through local communities [7].

Since Vietnam shares a long northern border with China and had previously experienced the severe acute respiratory syndrome (SARS) epidemic in 2003 [8], it recognized COVID-19 a threat from the outset, Vietnam first halt all flights to and from Wuhan, began rapid identification and testing of suspected cases, treatment for positives and instituted rigorous quarantine policies [9]. Unlike other countries like Germany only documented those infected and their direct contacts [10], Vietnam employed a four-tier approach to contact tracing and isolation. Tier one included confirmed cases who were isolated and treated in health facilities. Tier two were those who had close contact with confirmed cases and were asked to undertake testing and government-run quarantine for 14 days. Those in Tier three had close contact with Tier two cases and were required to self-isolate at their residential locations and under active monitoring and Tier four included the isolation of entire communities [11].

Beside the control measures, Vietnam government timely and constantly updated people the outbreak [12]. The Ministry of Health actively coordinated with media to conduct an intensive communication campaign raising public awareness of the COVID-19, promoting adoption of preventive behaviors (wearing masks, hand washing) and social distancing (e.g. stay home and avoid large group gathering) through national TV, loudspeakers, mobile communication motorbikes, street posters, the press and social media [5,9].

Despite limited resources, Vietnam’s early responses and decisive measures have kept the incidence of Covid-19 low thus far. There have been no new locally transmitted cases of COVID-19 in 41 days as of May 27, 2020. All reported 59 COVID-19 patients since April 16, 2020 were acquired from overseas and were detected while they were on a mandatory 14-day quarantine in the designated locations upon their arrival in Vietnam [5].

The purpose of this research paper is to examine and compare epidemiological characteristics of COVID-19 patients in Vietnam. Data from this study contribute to the epidemiology of COVID-19 in Vietnam and worldwide.

## Method

### Data sources

The data from this paper was obtained using publicly available information from the official website of the Vietnam Ministry of Health [5] during the period of 01/23/2020 to 05/27/2020 and was entered and organized using SPSS 22.0 (IBM Corp., Armonk, NY). The data provide details of each patient including date of diagnosis, age, sex, place of residence, travel and contact history and discharge status. Clinical data for each patient was not provided, however, the data summarized major symptoms of all COVI-19 patients. This study was conducted using public datasets and did not involve human subject and therefore did not require IRB approval.

### Analysis

Data were analyzed to describe the characteristics of the patients using mean and percentages. T-test, Chi-square test and Fisher’s Exact test were used to compare patients’ gender, age and regions of residence between two treatment status groups and between overseas and non-overseas travel groups. All analyses were conducted using SPSS 22.0.

### Results

A total of 327 patients (corresponding to the cumulative incidence of 34 per 10 million people) tested positive for COVID-19 and were hospitalized. The epidemiologic curse during the covering period is shown in Figure 1. A total of 278 (85.5%) recovered and were discharged from hospitals. The median age of all patients was 30 years old (range: 3 months-88 years old). The majority (83%) were 50 years old or younger. Fifty percent (n=164) were female (Table 1).

**Figure 1.**
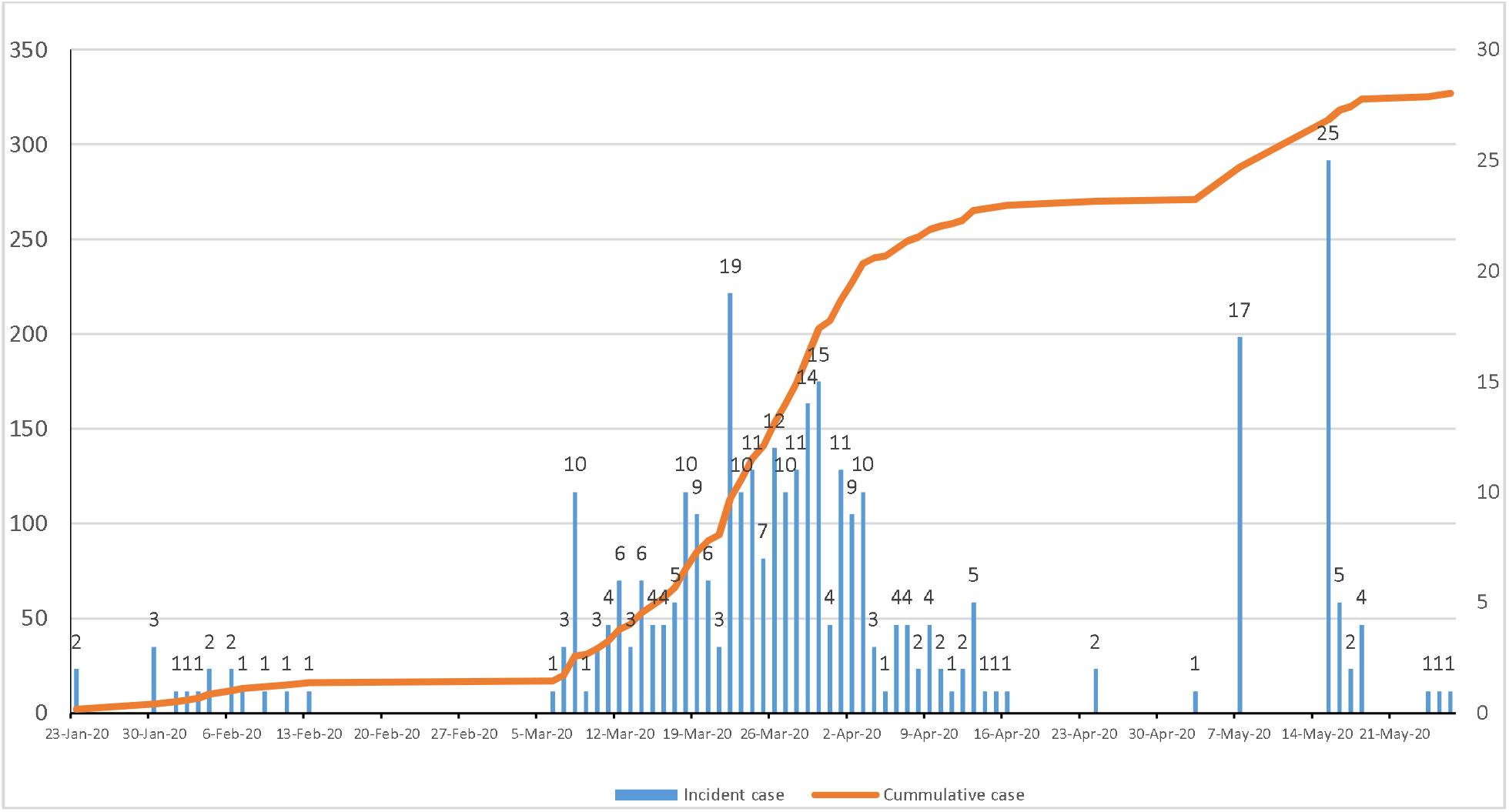
Incidence and cumulative cases of COVID-19 in Vietnam from 1/23/2020 to 05/27/2020

**Table 1.** Characteristics of patients with COVID-19 (n=327)

|  | No. (%) |
| --- | --- |
| **Age (Mean, range)** | (30, 3 months–88 years old) |
| <=10 | 7 (2.1) |
| 11-20 | 38 (11.6) |
| 21-30 | 98 (36.6) |
| 31-40 | 42 (15.7) |
| 41-50 | 40 (14.9) |
| 51-60 | 31 (11.6) |
| >60 | 28 (10.4) |
| **Sex** |  |
| Male | 123 (49.9) |
| Female | 145 (50.2) |
| **Region** |  |
| North | 208 (63.6) |
| Central | 24 (7.3) |
| South | 95 (29.1) |
| **Oversea travel history** |  |
| Yes | 223 (68.2) |
| No | 104 (31.8) |
| **Treatment status** |  |
| Under treatment | 49 (15.0) |
| Discharged | 278 (85.0) |

| **Nationality** |  |
| --- | --- |
| Vietnamese | 276 (84.4) |
| United Kingdom | 21 (6.4) |
| Brazil | 6 (1.8) |
| American | 5 (1.5) |
| Others | 19 (11.6) |

Note: Data as of May 27, 2020

A majority those who were infected inside the country was women (66.3%). Sixty eight percent of COVID-19 patients including students, workers and foreign visitors (n=223) had acquired the disease outside the country while 104 cases (31.6%) were infected within the country. Nearly 85.0% of the patients were Vietnamese while the remainder (15.6%) were foreigners including citizens of the United Kingdom, USA, Germany, Brazil and others.

Of 63 provinces, 31 reported having COVID-19 cases. Hanoi, the capital of Vietnam, had the highest number of patients (n=114), followed by Ho Chi Minh City (n=59), Thai Binh (n=30), Bac Lieu (n=21), Vinh Phuc (n=19) and the remaining had 1 to 13 cases (Figure 2). The North Region had the highest number of cases with 63.6% (n=106), followed by the South with 29.1 % (n=95) and the Central had lowest with 7.3% (n=24).

**Figure 2.**
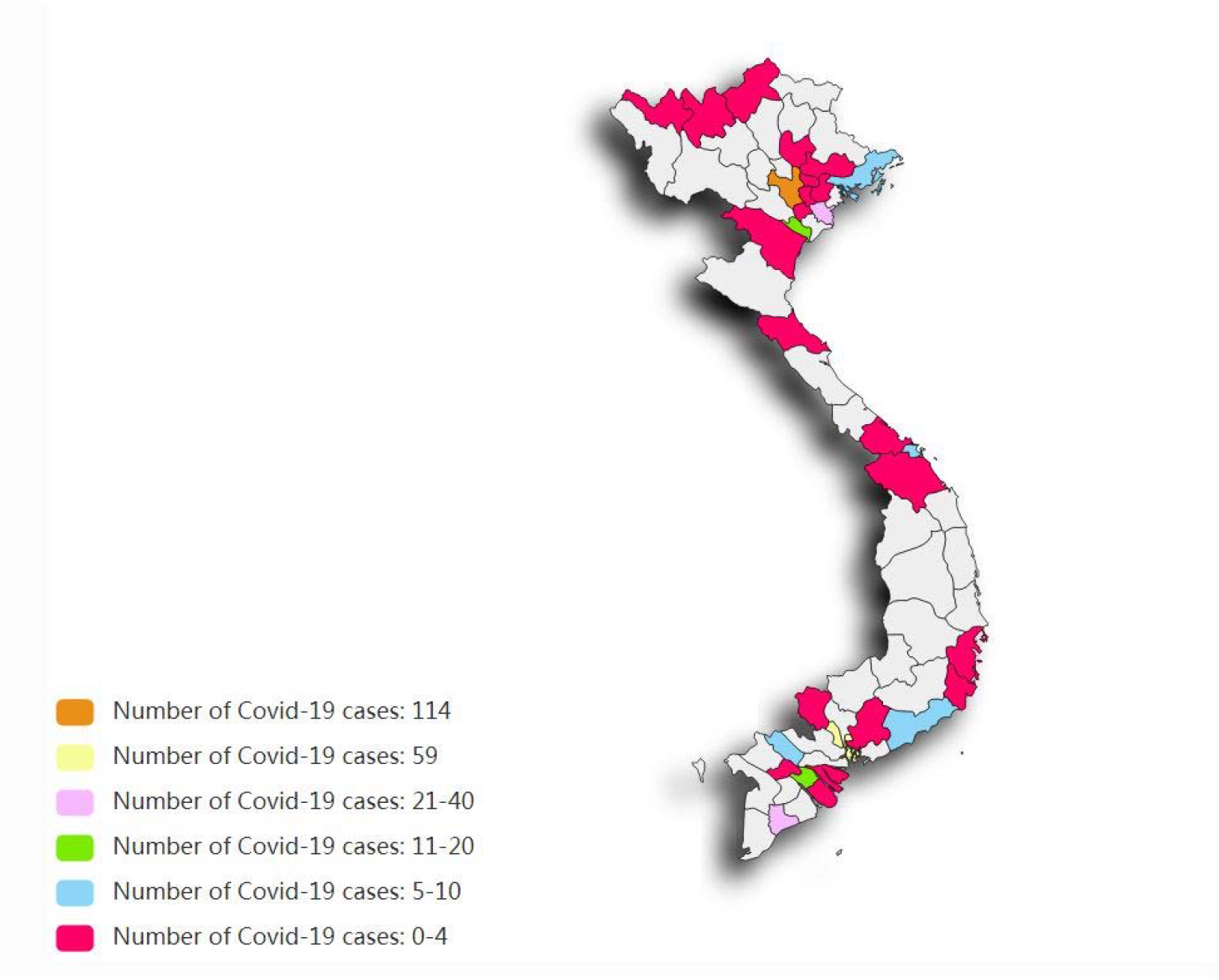
Frequency of COVID-19 cases in each province in Vietnam as of 05/27/2020

The results showed men were more likely to infected with COVID-19 from overseas travel [χ^2^(1, n = 327) = 15.27; p<0.001] (Table 2). There were significant differences in age between overseas travel (M=33.1, SD=14.8) and non-overseas travel (M=39.3, SD=15.2) with younger patients being associated with international travel; [t(325) =3.4, *p* =.001]. Also, there were significant differences between regions based on patients ‘overseas travel history. Compared to those in the North (64.9%) and Central (58.3%), patients in the South (77.9%) reported the highest level of overseas travel [χ^2^(2, n= 327)=6.83; p=0.044].

**Table 2.** Factors associated with oversea travel history of COVID-19 patients in Vietnam

| Characteristics | Overseas travel history |  | T-test or Chi-square test |
| --- | --- | --- | --- |
|  | Yes | No |  |
| **Age** (mean, SD) | 33.1 (14.8) | 39.3 (15.2) | $t(325)=3.4, p=.001$ |
| **Gender** | | | $\chi^2(1, n = 327)=15.27; p<0.001$ |
| Female | 95 (57.9%) | 69 (42.1%) |  |
| Male | 128 (78.5%) | 35 (21.5) |  |
| **Regions** | | | $\chi^2(2, n= 327)=6.83; p=0.044$ |
| North | 135(69.4%) | 73 (35.1%) |  |
| Central | 14 (53.8%) | 10 (41.7%) |  |
| South | 74 (77.9%) | 21 (22.1%) |  |

In terms of treatment status, women (54.7%) had a higher rate of discharge compared to men (45.3%) [χ^2^(1, n = 327)=15.27; p<0.001] (Table 3). Patients infected with COVID-19 inside the country were more likely to be discharged from the hospital than those infected from overseas (p<0.001). There were significant differences between regions of residence based treatment status (p=0.028) with patients in the Central (100%) reporting the highest level of being discharged. No significant differences were found in age between those who have been recovered and discharged and those who are still treated in the hospital (p=0.258).

**Table 3.**
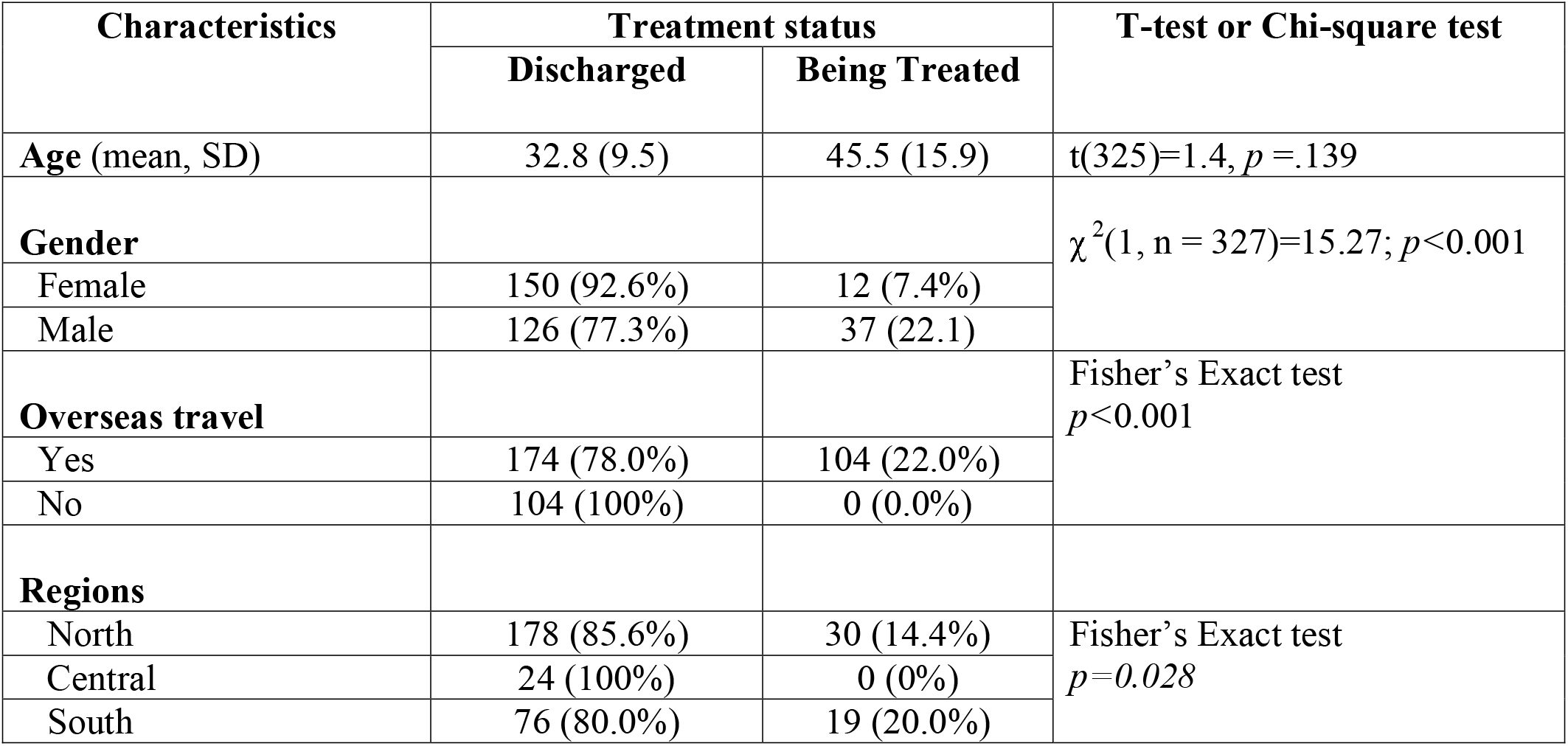
Factors associated with treatment status of COVID-19 patient in Vietnam (n=327)

Only 36.5% of patients had the symptoms when confirmed positive to the virus. Among symptomatic patients, common symptoms included 22.8% fever, 18.5% cough, 9.9 % sore throat and only 3.4% shortness of breath.

## Discussion

While most of the countries including China, Italy, South Korea and US reported fatalities from COVID-19 [3], there have been no reported fatalities among the patients in Vietnam. Nearly 86% of the patients have recovered and been discharged from the hospital. Other Asian countries such as Hong Kong, and Taiwan, which had experiences in handling and dealing with the severe acute respiratory syndrome (SARS) epidemic in 2003 and were in a good position responding to COVID-19, still suffered from COVID-19 fatalities [13,14], the fact Vietnam has no COVID-19 related deaths is an extraordinary outcomes and deserve further research on its handling and treatment of COVID-19 patients.

The finding revealed that a majority of the infection cases in Vietnam occurred among the people below 60 years old. This was in contrast with the infection rates of COVID-19 in other countries like Iran [15], France [16] and Italy [17] in which the infection rates were more dominant among the people over 60 years old.

Unlike other countries (e.g. the USA) [18], where asymptomatic people are recommended to stay home and are not recommended to get tested for COVID-19, in Vietnam home-based isolation of confirmed cases is not allowed and all COVID-19 asymptomatic cases were hospitalized. Evidence has shown that COVID-19 can transmit while pre-symptomatic and asymptomatic [19]. Therefore, the transmission situation in Vietnam could have been worse if these asymptomatic individuals were not detected early and hospitalized as they could have silently spread the virus in their community.

The majority of infection cases including foreign tourists, students and workers coming back from oversea were acquired from abroad. The government’s decision to aggressively screen those coming to the country and quarantine them for 14 days in the concentrated isolation locations as soon as they came to Vietnam helped prevent the further transmission.

Over 66% of women were infected with COVID-19 from an index patient inside the country. It could be that women are more likely to be caregivers for the original COVID-19 infected people and hence they were more likely to be infected. Other possibilities include a lack of personal protection equipment and lack of awareness of COVID-19. Further exploratory research is needed to explore the gender disparities of COVID-19 infection in Vietnam.

This study should be interpreted in light of certain limitations. While there is no case fatality yet, 49 patients remain in the hospital and outcomes yet to be known. A time of this writing, Vietnam has not reported any locally transmitted COVID-19 cases for 41-days. All new cases registered since April 16, 2020 are Vietnamese workers and students returning from overseas. However, the pandemic is still going on worldwide, the data therefore should be read as time-dependent and temporary. At this time, no serological data is available on antibody responses in the population.

## Conclusions

The findings showed that a majority of COVID-19 cases in Vietnam were acquired overseas. A significantly higher number of women than men were infected inside the country calling for further research about gender disparities in the fight against COVID-19 in Vietnam. Although the pandemic continues to spread around the world, Vietnam has not reported a locally transmitted infection case in 41 days, which might suggest that Vietnam has kept the COVID-19 outbreak under control. Vietnam serve as a standard for comparative studies of COVID-19 containment and mitigation. However, the country still needs to remain vigilant as the pandemic is still evolving.

## Data Availability

The data was obtained from the publicly available information of the official website of Vietnam Ministry of Health covering a period of 01/23/2020 to 05/27/2020

## Acknowledgements

The authors would like to acknowledge Dr. Audrey Chapman for her helpful comments and suggestions on this paper.

## Author contributions

Author contributions:

Study concept and design: Ha; Data acquisition and statistical analyses: Ha; Interpretation of data: Ha, Lewis and Ruaño; Drafting of manuscript: Ha; Critical revision of the manuscript for important intellectual content: Ha, Lewis and Ruaño.

